# The cross-disciplinary integration of acupuncture and modern medicine: an analysis from the perspective of subject categories

**DOI:** 10.1101/2022.11.27.22282799

**Authors:** Xin Zhang, Zhiguang Duan

## Abstract

The cross-disciplinary integration of acupuncture with modern medicine is an important guarantee to maintain its advantages. Based on 13896 acupuncture articles and 404726 references in the Web of Science Core Database, this paper analyzes the cross-disciplinary integration of acupuncture and modern medicine. It was found that the trend of cross-disciplinary integration between acupuncture and modern medicine is more and more obvious. Neurosciences, Anesthesiology, Oncology, Obstetrics & Gynecology, Psychiatry, and Gastroenterology & Hepatology are the subjects with the most crossover and integration, accounting for 56% of the total disciplines. At the same time, more studies have begun to use modern medical methods to study the mechanism of acupuncture. Biochemistry & Molecular Biology, Pharmacology & Pharmacy, and Medicine, Research & Experimental have been major areas of interdisciplinary collaboration in recent years. China, the United States, and South Korea are the three countries with the largest number of publications. Among them, Chinese scholars pay more attention to the role of acupuncture in the treatment of neurological diseases, Korean scholars pay more attention to drug acupuncture, especially bee venom acupuncture, and American scholars pay more attention to the role of acupuncture in pain management. However, acupuncture has less cross-disciplinary cooperation with heart and cardiovascular system, peripheral vascular disease, and even less with non-medical disciplines. Using modern medical technology and methods to carry out research on the mechanism of acupuncture and moxibustion and strengthen the cooperation with informatics, statistics, physics and other disciplines may be an important direction for the modernization of acupuncture.

关键词: 针灸; 跨学科; 学科范围

## Introduction

With the development and improvement of medical technology, previously incurable diseases have been solved, previously helpless infectious diseases have been controlled, and life expectancy has increased. However, modern medicine is not omnipotent and has some drawbacks. For example, modern medicine mainly uses opioids in the treatment of pain, but they are addictive^1^. Thrombolytic drugs require a time window in the treatment of stroke, and the incidence of disability is higher after the time window^2^. Radiation therapy for cancer may have side effects such as nausea, vomiting, and fatigue^3^. As an important part of Traditional Chinese medicine, acupuncture has been developed in China for thousands of years and has an obvious advantage in making up for the shortcomings of modern medical treatment^4^. Over the last few years, researchers around the world have managed and did a large number of studies on the efficacy and mechanism of acupuncture. Studies have shown that acupuncture has a good analgesic effect and will not cause liver and kidney damage, gastrointestinal diseases, and other side effects^5^. Maria et al. confirmed that electroacupuncture has a good effect on analgesia during tooth extraction^6^. Li et al. believed that acupuncture had better efficacy and safety in the treatment of dysphagia after stroke compared with drug therapy^7^. Kim et al. believed that moxibustion could effectively treat cancer-related fatigue^8^. These studies have proved that acupuncture has a good curative effect.

Therapy acupuncture has not been fully accepted by modern medicine. Acupuncture can achieve innovative development only by absorbing, drawing on, and integrating the knowledge of other disciplines. In the field of academic research, interdisciplinary integration is directly reflected in the publication of interdisciplinary research papers^9^. As an important embodiment of knowledge sources, the subject types of references represent interdisciplinary situations. Therefore, it is very important to understand the subject distribution of acupuncture literature and reference literature to promote the innovation and development of acupuncture.

This paper takes the Web of Science Category(WC) of acupuncture articles and their references as the research object and adopts bibliometrics to understand the crossover and integration of acupuncture and modern medicine. DIV was used to measure the degree of intersection between acupuncture and other disciplines. To understand the current development of acupuncture and provide a reference for its later development.

## Methods

### 1. Data sources and search strategies

We conducted a comprehensive online search on August 10, 2021, using the Web of Science (WOS) core database. The search terms were acupuncture. The retrieval period is not limited.

### 2. Criteria for inclusion and dispatch of documents

Two independent reviewers sifted through the document. Document related to acupuncture was included, and duplicate publications, letter, meeting abstract, and news items were excluded.

### 3. Data analysis and Visualization

Excel2016 was used to sort out Web of Science Category(WC), keywords, and other information.

### 4. Interdisciplinary measurement index

Loet Leydesdorff combined three indicators of interdisciplinary Variety, Balance, and Disparity, and proposes a new DIV to measure interdisciplinary^10^.

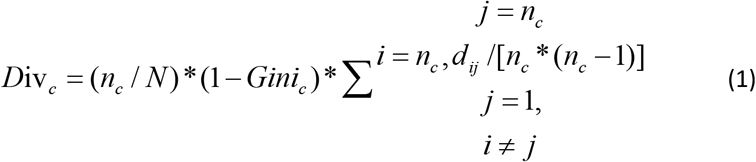

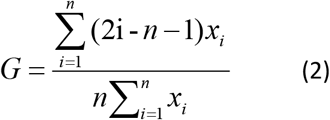

In Formula (1), n_c_/N stands for “Variety”, n_c_ stands for the number of citation WC in Article C, and N stands for the number of all citation WC. The “balance” can be manipulated using the Gini coefficient. Where x is an observed value of references WC, n is the number of value observed, and i is the rank of values in rising order. Disparity is a revision of RS Diversity. “d_ij_” is the difference between the WC_i_ and the WC_j_, expressed by cosine.

## Results

### 1. Search results

14034 documents and 586024 references were retrieved from WOS. A total of 138 theme documents without references and 181298 references without WC were excluded. The document retrieval flowchart is provided in Figure 1.

**Fig. 1.**
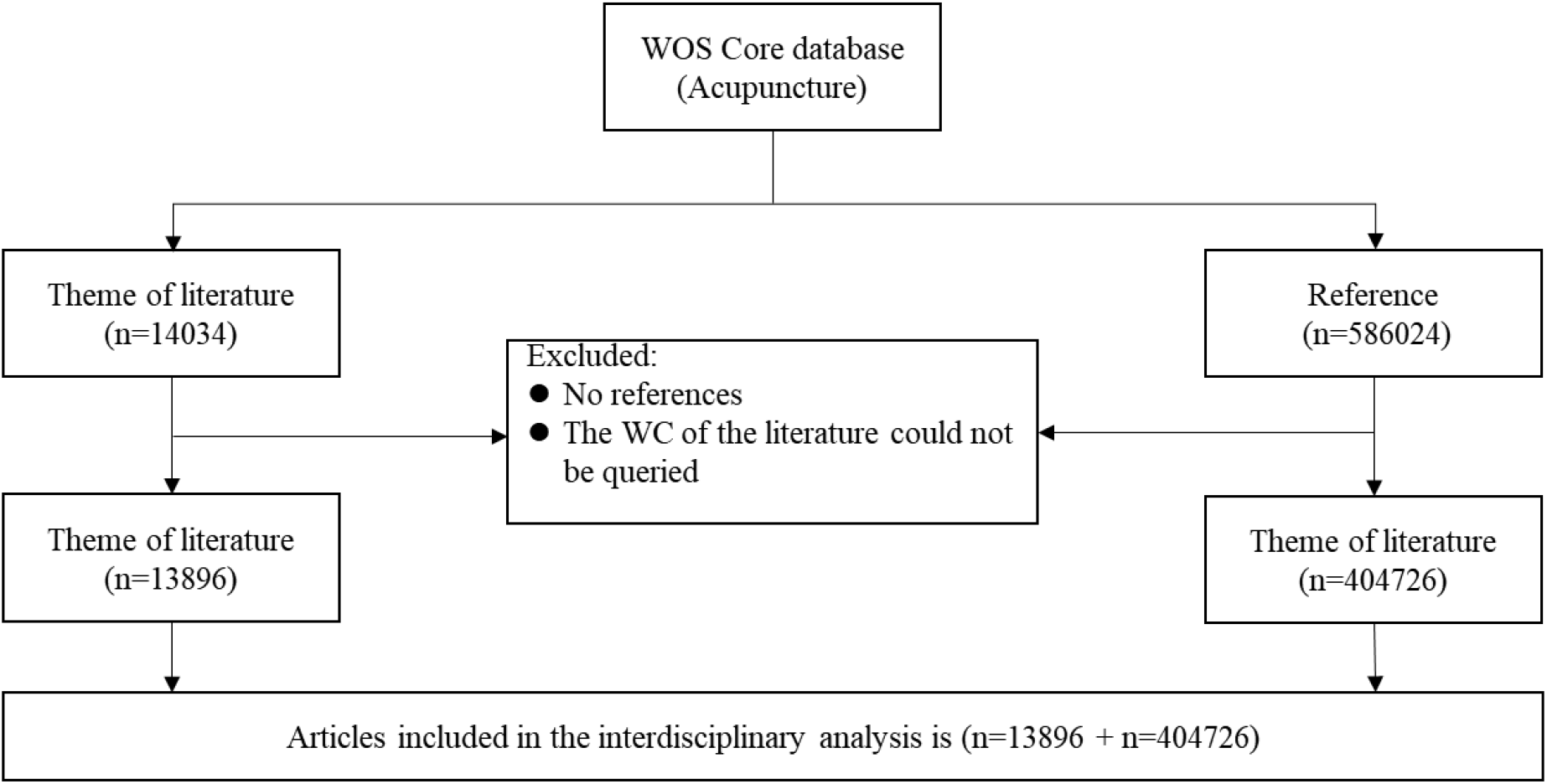
Retrieval Flowchart

### 2. The WC of the subject articles

The first article on acupuncture appeared in 1973. The number of relevant articles before 1999 is relatively small. In 1999, there was a sudden increase in the number of publications, about five times that of 1998. From 1999 to 2020, the number of articles showed a popular thing.

At least one WC will be assigned to every journal in WOS. When an article has two or more WC, we call it a multidisciplinary article, and conversely, we call it a single-disciplinary article. The number of multi-disciplinary articles continued to rise in 1999 and 2000.

Table 1 shows the top 10 articles WC and multidisciplinary articles WC. A total of 143 WC were involved in 13896 articles, and the top 20 WC accounted for 79.1% of the total WC. Add up to 136 WCS were involved in 3631 multidisciplinary articles, and the top 20 WCS accounted for 72.8%. The WC of the two types of articles is 78.3 the same.

**Table 1.** Top 10 published articles WC and multidisciplinary articles WC

| Number | Multidisciplinary articles WC | Frequency | All articles WC | Frequency |
| --- | --- | --- | --- | --- |
| 1 | Neurosciences | 1124 | Integrative & Complementary Medicine | 4307 |
| 2 | Clinical Neurology | 782 | Medicine, General & Internal | 1850 |
| 3 | Integrative & Complementary Medicine | 685 | Neurosciences | 1529 |
| 4 | Medicine, General & Internal | 534 | Clinical Neurology | 965 |
| 5 | Rehabilitation | 370 | Medicine, Research & Experimental | 666 |
| 6 | Pharmacology & Pharmacy | 309 | Anesthesiology | 444 |
| 7 | Anesthesiology | 303 | Oncology | 408 |
| 8 | Medicine, Research & Experimental | 260 | Pharmacology & Pharmacy | 369 |
| 9 | Oncology | 247 | Obstetrics & Gynecology | 362 |
| 10 | Cell Biology | 243 | Rehabilitation | 361 |

### 2. The WC of the References

From 1999 to 2020, the number of references per article and the number of reference WC increased year by year. The average number of references in 2020 was 34.4, involving 165 WC. Figure 2 shows the top 18 WC in terms of quantity, accounting for 70.2% of the total WC in the references. However, Reproductive Biology, Dermatology, Allergy, Otorhinolaryngology, Respiratory System, and Ophthalmology accounted for less than 1% of the total WC in the references.

**Fig. 2.**
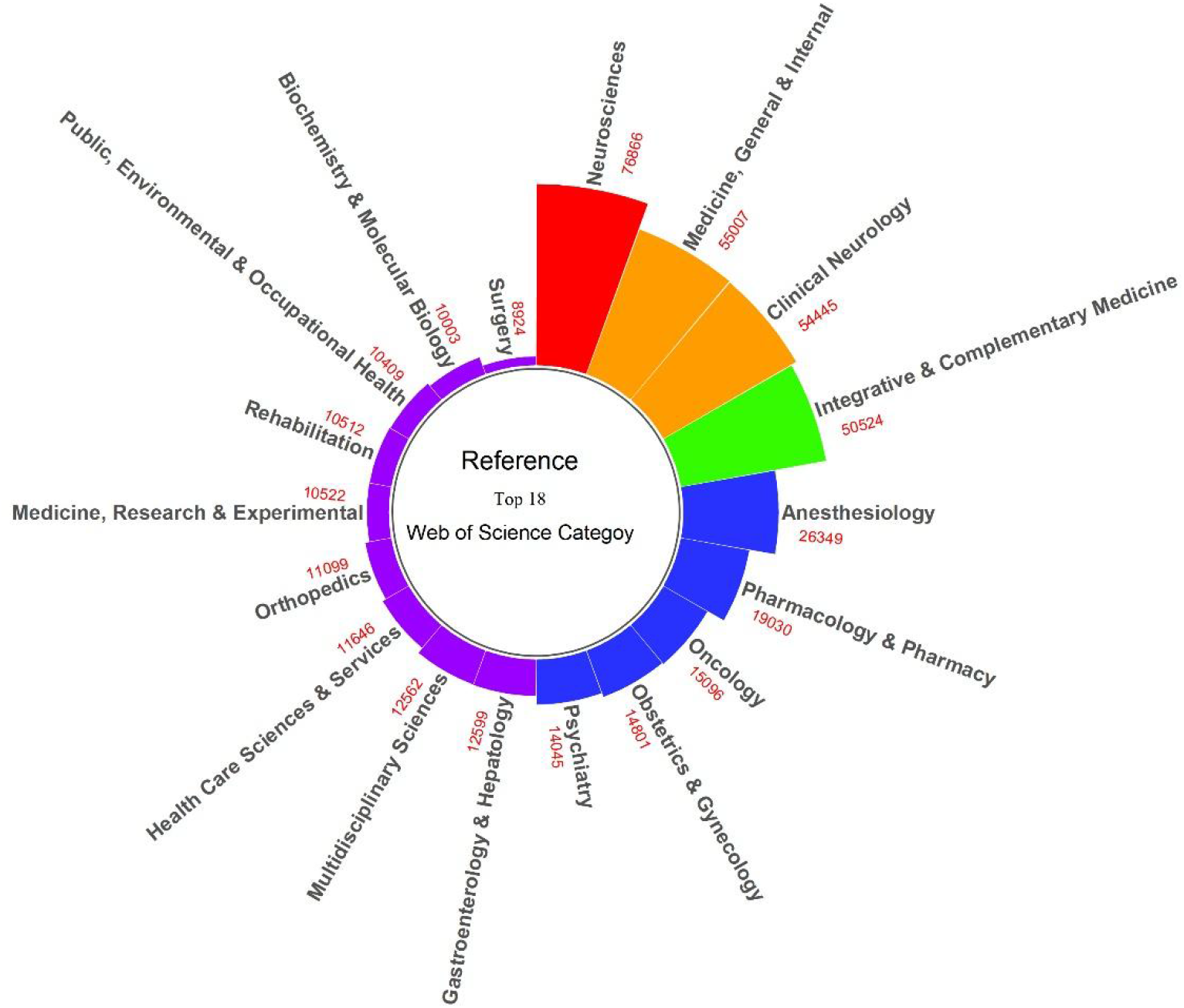
Top 18 WC in the references. The height of the bar in the figure represents the frequency of WC.

Figure 3 shows the WC changes of the top 18 in number from 1999 to 2021.. Except for Obstetrics & Gynecology, which has declined since 2015, all the other 17 WC categories have shown an upward trend. Medicine, Research & Experimental, Biochemistry & Molecular Biology, Multidisciplinary Sciences, Gastroenterology and Hepatology is growing rapidly in recent years, and the number of Anesthesiology grow slower.

**Fig. 3.**
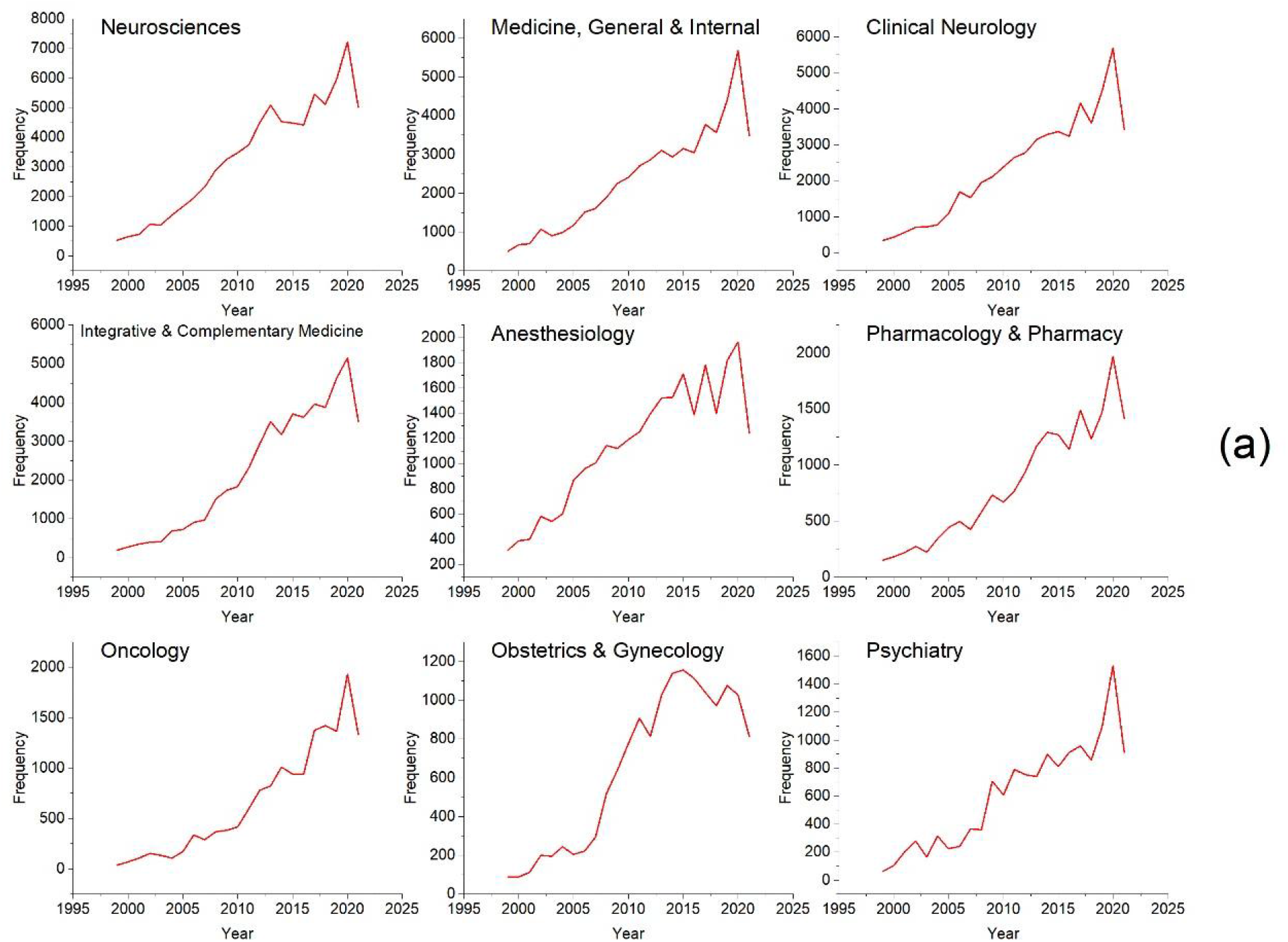
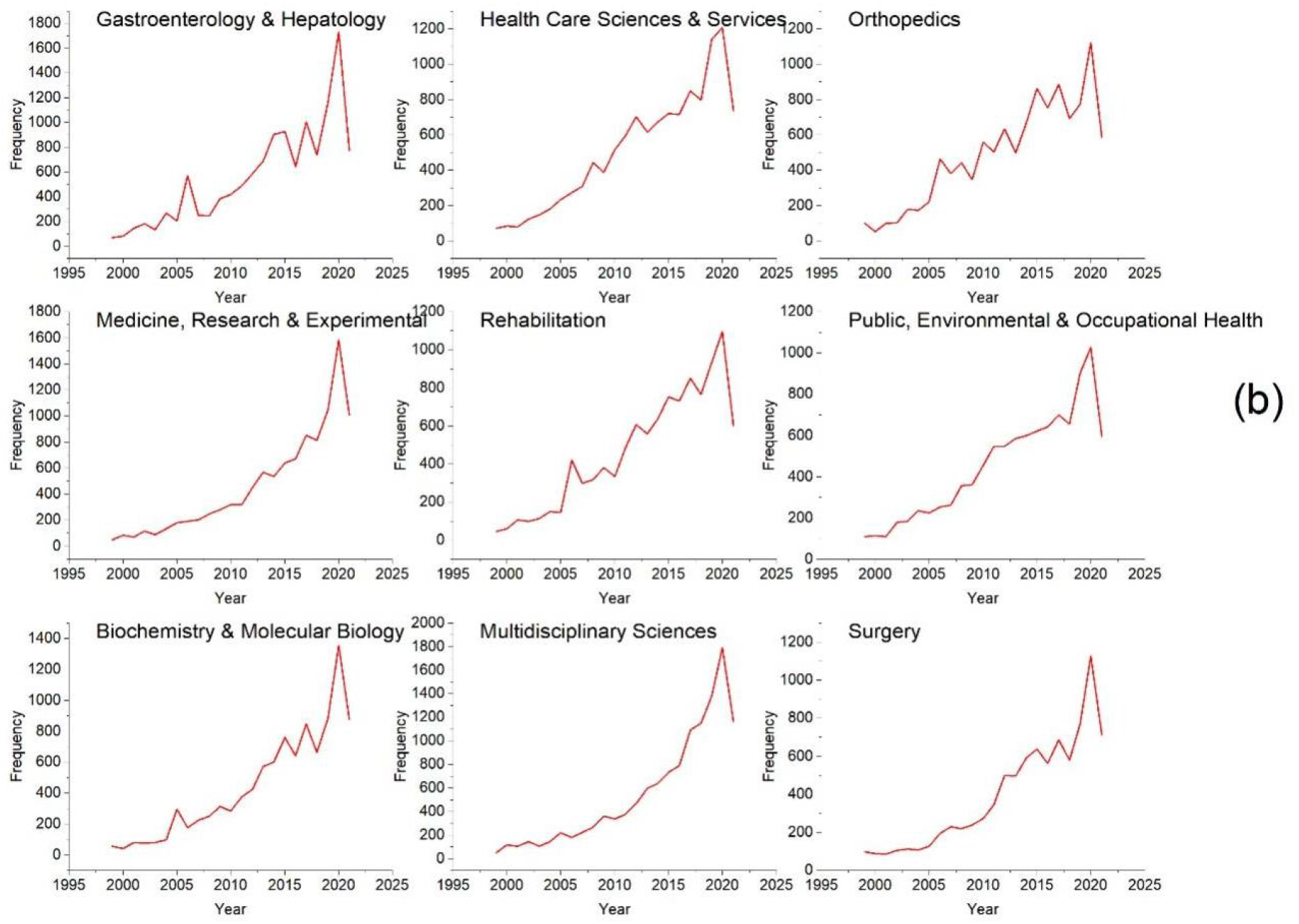
The number change of WC in the top 18 from 1999 to 2021. (a)Neurosciences, Medicine, General & Internal, Clinical Neurology Integrative & Complementary Medicine, Anesthesiology, Pharmacology & Pharmacy, Oncology, Obstetrics & Gynecology, Psychiatry (b) Gastroenterology & Hepatology, Health Care Sciences & Services, Orthopedics, Medicine, Research & Experimental, Rehabilitation, Public, Environmental & Occupational Health, Biochemistry & Molecular Biology, Multidisciplinary Sciences, Surgery

### 3. DIV is used to evaluate the interdisciplinarity of the article

Figure 4 is the trend chart of DIV over time. As can be seen from the figure, the DIV of acupuncture articles in recent years has been continuously increasing. China, the United States, and South Korea are the top three countries in terms of the number of articles published, among which China has published 4,263 articles, the United States has published 2,759 articles, and South Korea has published 1,273 articles. China, the United States, and South Korea accounted for 56.2% of the total publications. DIV of the United States, China, and Korea all showed an upward trend, but DIV of the United States was higher than that of China and Korea.

**Fig. 4.**
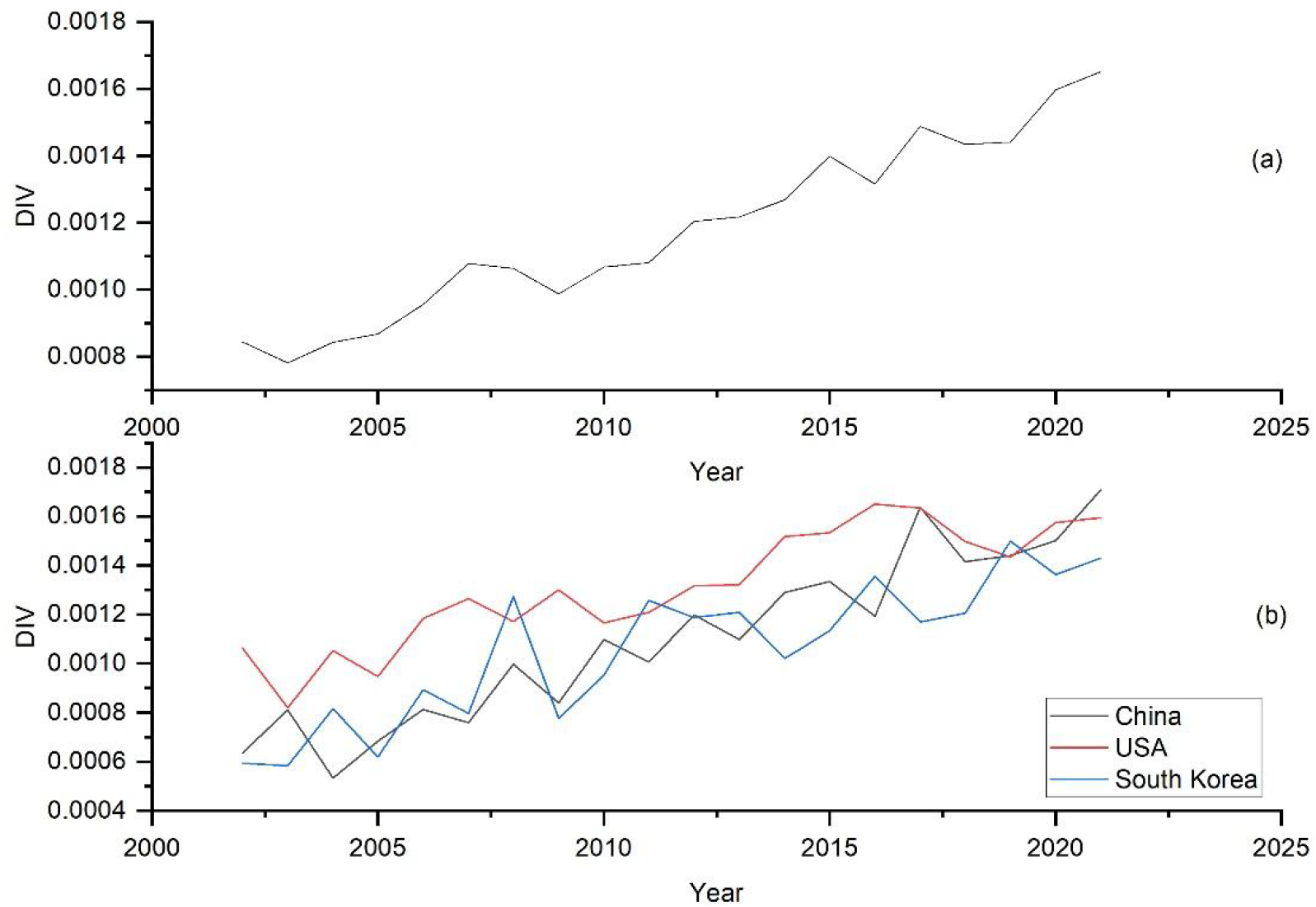
The trend chart of DIV. (a) The number trend of articles of different DIV groups. (b) DIV change trend chart of China, USA and South Korea in different years.

Arrange the DIV of 13,896 articles in descending order. According to the quartile spacing, the top 25% of articles were divided into the High-DIV group, the last 25% into the Low-DIV group, and the middle 50% into the Middle-DIV group. Among the top 10 WCs, 80% of High DIV groups are the same as Middle DIV groups, and 60% are the same as Low DIV groups..

## Discussion

According to the 13,896 acupuncture articles, the number of published papers and the types of WC in the articles is increasing year by year, but WC mainly focuses on Integrative & Complementary Medicine and Medicine, General & Internal. This may be related to the different understanding of acupuncture in different countries. In China, acupuncture has been developed for thousands of years, has its own independent theoretical system, has been proven to have distinct clinical efficacy, and is an important part of Traditional Chinese medicine^11^. In South Korea, acupuncture is regarded as a kind of Oriental medicine with an ancient history. After many years of development, acupuncture therapy with its characteristics has been formed^12^. But in the United States, acupuncture is not included in the mainstream medical community but is defined as a medical intervention^13^. In “Multidisciplinary Article”, WC is mainly Neurosciences and Clinical Neurology, suggesting that acupuncture is now the most widely used in nerve-based disease. Taking cerebrovascular disease as an example, the most effective treatment means are thrombolysis and intervention, but both of these methods are limited by time window. How to help patients recover neurological function in the follow-up treatment process, more treatment means are needed^14^. Chinese acupuncture has shown remarkable results in treating stroke. Several RCT studies in the United States have also proved that acupuncture has a good effect on brain health^15^. Acupuncture has also been shown to be effective in the treatment of pain associated with neurological diseases. Pain is one of the hardest signs for Parkinson’s patients, and studies have shown that acupuncture can effectively reduce pain perception^16^.

According to 404,726 references, the number and types of WC are increasing, pointing to that the width of knowledge sources for acupuncture research is expanding. As can be seen from the top 18 references WC in the past decade, the number of Medicine, Research & Experimental, Biochemistry & Molecular Biology, and Pharmacology & Pharmacy was increased. For example, Zheng^17^ explored the rule of acupuncture analgesia through TRPV4. Ding^18^ studied acupuncture in the treatment of spinal cord injury by controlling the Shh/GLI-1 Signaling Pathway. Chanya Inprasit conducted a study on the reduction of Complete Freund’s Adjuvant pain caused by electroacupuncture by regulating TRPV1^19^. According to HU^20^ research, acupuncture and Chinese medicine combined with stress have a good effect with fewer side effects. Pain, nausea, vomiting, fatigue, and other adverse reactions often occur during the treatment of cancer patients, which are almost unavoidable in current medical treatment. Acupuncture is also widely used in cancer treatment as an alternative therapy with very few side effects^21^. Recent studies have shown that electroacupuncture can reduce the cytokines, Iba1 and GEAP, thus have analgesic effects on pulp injury^22^. Other studies have summarized five mechanisms by which acupuncture can benefit stroke rehabilitation^23^. Different from the middle DIV-group and the Low-DIV group, the High-DIV group has a large amount of knowledge input in Medicine, Research & Experimental, Biochemistry & Molecular Biology, and Pharmacology & Pharmacy, which also proves that modern medical research methods are constantly applied in the research of acupuncture.

Reproductive Biology, Dermatology, Allergy, Otorhinolaryngology, Respiratory System, and Ophthalmology are less than 1%. It shows that acupuncture and moxibustion were not well integrated with these subjects. However, acupuncture and moxibustion have a good effect in the treatment of ophthalmology, respiratory system, and ent diseases. Take acupuncture as an example in the treatment of myopia. In recent years, myopia has developed rapidly in East Asian countries, especially among school-age children^24^. Studies have shown that the prevalence rate of myopia among 17-18 Chinese children is 70-85%^25^. If we do not control myopia and allow it to progress, it may lead to macular detachment, retinal detachment, and even blindness, which seriously endangers the health of teenagers. Compared with outdoor activities, corneal orthosis, and surgical intervention, acupuncture has the advantages of no time limit, no age limit, and large-scale popularization in the prevention and treatment of myopia. Huang et al.^26^conducted a RCT study on acupuncture in the treatment of myopia, proving that acupuncture can effectively improve myopia. Katarzyna et al.^27^ discussed the acupuncture points for the treatment of myopia. Acupuncture is also effective in treating other diseases, and further research is also needed.

China, the United States, and South Korea are the three countries with the largest amount of publications. Chinese researchers have focused on neurological diseases such as stroke, nerve regeneration, Neuropathic pain and Alzheimer’s disease. According to the stroke treatment goal set by the American Heart Association/American Stroke Association, the time management of thrombolysis within 60 minutes is ≥ 75%, and the rate of meeting the standard is 51% in the United States, while the rate of meeting the standard is only 9% in China^28^. Moreover, the incidence of disability after discharge is 25% to 35% in those who do not meet the standard. Several trials have shown that acupuncture can help stroke patients recover and improve their quality of life^29^. The South Korean researchers focused on Pharmacopuncture,, specifically bee venom acupuncture. Multiple RCT tests have demonstrated that bee venom acupuncture is effective in relieving pain and treating arthritis^30^. The USA researchers focused more on acupuncture’s role in pain treatment. According to statistics, more than 100 million people in the United States suffer from chronic pain^31^. Currently, opioids are the treatment for pain. In 2018, the United States passed the pain and opioid addiction law to prevent and treat opioid addiction. Acupuncture has the advantages of fewer side effects and no addiction in pain treatment and has been proven to be effective in treating pain in multiple RCTs^32^.

## Conclusion

In a word, the interdisciplinary cooperation between acupuncture and modern medicine is strengthening, but there are still few studies on the mechanism of acupuncture using modern medicine techniques and methods, and even less with other non-medical disciplines. Therefore, the use of modern medical technology and methods to research the mechanism of acupuncture, and strengthen the cooperation with informatics, statistics, physics, and other disciplines may be an important direction for the modernization of acupuncture.

## Data Availability

All data produced in the present study are available upon reasonable request to the authors

